# Surgical, Anesthesia and Obstetrics (SAO) workforce production capacity in India: A retrospective analysis of postgraduate and sub-specialty training spots

**DOI:** 10.1101/2023.01.12.23284480

**Authors:** Shirish Rao, Siddhesh Zadey

**Author notes:** **Correspondence:** Siddhesh Zadey, BSMS MSc-GH; ASAR Office Address: D2 Sai Heritage, New DP Road, Aundh, Pune, Maharashtra, India 411007.

## Abstract

**Objective:** To assess the SAO production capacity of India i.e., the number of postgraduate (PG) and sub-specialty (SS) surgical training spots per 10 million population across multiple specialties and subspecialties at national and state levels.

**Methods:** A retrospective secondary data analysis of PG and SS SAO spots across 36 states for 2018 using data from the National Health Profile (2019) was conducted. The mid-year 2018 populations were obtained from the census-based population projections. The number of PG & SS SAO spots per 10 million population were calculated and divided based on the type of program (diploma, MD/MS & DM/MCh) and type of SAO specialty to investigate SAO workforce production capacity in each state. Ratios of PG spots per 100 MBBS spots and SS spots per 100 PG spots were also calculated. Data was wrangled using Google Sheets, analyzed in JASP v0.16.0.0, and visualized using Datawrapper.

**Results:** There are a total 13793 PG and 1350 SS SAO spots leading to densities of 104.60 and 9.90 per 10 million people, respectively. PG spot density for General Surgery (23.56), Anesthesia (24.81), and OBGYN (21.55) were comparable and were much higher than Orthopedics (14.64), Ophthalmology (11.96), and Otorhinolaryngology (8.08). SS spot densities were greater for Urology (1.90), Neurosurgery (1.86), and Cardiothoracic and Vascular Surgery (1.83), followed by Plastic Surgery (1.52), and Pediatric Surgery (1.27). The average density was significantly lower for PG SAO specialties than non-SAO specialties (p=0.001) whereas there was no significant difference in densities for SS SAO and non-SAO specialties (p=0.197). For 100 MBBS spots, there were only 20 PG SAO spots while for 100 PG SAO spots, there were just 9 SS SAO spots available. The distribution of spots was geographically uneven with about two-thirds of spots concentrated in 10 states/union territories.

**Conclusion:** Annually, India can produce only about 24 General Surgeons, 15 Orthopedic Surgeons, 12 Ophthalmologists, 8 Otorhinolaryngologists, 25 Anesthetists, 22 OBGYNs, and 10 sub-specialist surgeons per 10 million population. The distribution of spots is inequitable across the states. Hence, scale up of surgical training capacity needs to be carried out with attention to reducing disparities.

**Highlights:**

- We present a novel pan-India analysis of SAO workforce production capacity.
- In 2018, India had 13793 postgraduate and 1350 subspecialty SAO training spots.
- 59 surgeons and 10 subspecialist surgeons are produced per 10 million people.
- Competitiveness of matching into residency-level programs is assessed.
- Every 100 MBBS spots had 20 PG SAO spots. Every 100 PG SAO spots had 9 SS SAO spots.

## Introduction

India faces a severe shortage of human resources for health. There are about 800,000 active allopathic doctors making the density to be 61 per 100,000 population much below the recommended density of 207.^1,2^ The shortage is also observed in the surgical, anesthesia, and obstetric (SAO) workforce. Based on the dataset compiled by the Lancet Commission on Global Surgery (LCoGS), in 2009, India had 31,560 surgeons, 20,280 anesthetists, and 29,310 obstetricians making up an SAO density of 6.5 per 100,000 population.^3^ An accompanying analysis demonstrated that India needed 291,824 more SAO personnel to meet the target density of 20 by 2030.^4^ This can be achieved by an increase in SAO production capacity.

Medico-surgical training in India is spread over multiple subsequent degree programs extending over 7.5-11.5 years. Allopathic doctors begin their training in a Bachelor of Medicine and Bachelor of Surgery (MBBS) degree program of 5.5 years including a year of clinical internship.^5^ This is followed by three years of post-graduation (PG) in the case of a Master of Surgery (MS) or Doctor of Medicine (MD) program or two years of PG in the case of equivalent diploma programs. Further subspecialization (SS) includes Master of Chirurgiae (MCh) or Doctorate of Medicine (DM) training that are offered as 3- or 6-year programs following MS/MD (not diploma) or MBBS, respectively.^6^ Candidates are matched into PG or SS programs through competitive exams including but not limited to National Eligibility cum Entrance Test (NEET)-PG or NEET-SS respectively.^7,8^ The structure, duration, competencies focused, and methods of evaluation in case of Indian surgical training present different strengths and challenges compared to training in high-income countries such as the United States or the United Kingdom. ^9^

While SAO production capacity is critical for determining SAO workforce scale-up, there is limited literature on the topic for India. Previously, a report sponsored by the World Health Organization (WHO) used data from the Medical Council of India (now replaced by the National Medical Council) to find that in 2015, India had 9037 SAO (General Surgery, Anesthesia, Obstetrics & Gynecology, Orthopedic Surgery, Otorhinolaryngology, and Ophthalmology) PG spots corresponding to a density of 75 training spots per 10 million population. This report further focused specifically on the southern socioeconomically developed state of Kerala to find 411 SAO PG spots with a density of 123 spots per 10 million population.^10^

A systematic nationally representative analysis of SAO workforce production capacity is missing. This study aimed to assess SAO workforce production capacity across multiple specialties and subspecialties at national and state levels.

## Methods

### Study Design and Data Sources

We conducted a retrospective analysis of secondary data. Data on PG and SS SAO training spots across 36 states and union territories as of December 2018 were acquired from the National Health Profile report of 2019.^11^ National and subnational census-based mid-year population projections for 2018 were taken from the 2019 report by the National Commission on Population.^12^

### Variables and Definitions

A postgraduate (PG) SAO spot was defined as a Diploma or Master in Surgery (MS) training spot which offered specialization in one of the following areas: General Surgery, Anesthesia, Obstetrics & Gynecology (OBGYN), Orthopedic Surgery, Otorhinolaryngology, and Ophthalmology. Non-SAO PG specialties included Internal Medicine, Chest Medicine, Dermatology, Radiology, Psychiatry, Community Medicine, Transfusion Medicine, Immunology, Aviation Medicine, Radiotherapy, Nuclear Medicine, Sports Medicine, Emergency Medicine, Pediatrics, Physical Rehabilitation Medicine, Anatomy, Physiology, Biochemistry, Pharmacology, Microbiology, Pathology, Forensic Medicine, Public Health, and Health Administration.

A subspecialty (SS) SAO spot was defined as a Magister Chirurgiae (MCh) training spot that offered specialization in one of the following areas after PG-level SAO training: Urology (or Genitourinary Surgery), Neurosurgery, Plastic Surgery, Cardiothoracic and Vascular Surgery (including spots specific to Vascular Surgery and Thoracic Surgery), Pediatric Surgery, Onco-surgery, Gastrointestinal Surgery, Endocrine Surgery, Head-Neck Surgery and Cardiac Anesthesia. Different programs namely Cardiothoracic Surgery, Thoracic Surgery, Vascular Surgery, and Cardiothoracic and Vascular Surgery are available as SS courses. However, we have considered all these programs under “Cardiothoracic and Vascular Surgery’’. The above-mentioned SS courses except Head and Neck Surgery and Cardiac Anesthesia can be pursued only after completing PG in General Surgery. Head and Neck Surgery can be pursued after General Surgery as well as Otorhinolaryngology while Cardiac Anesthesia can be pursued after PG in Anesthesia. Non-SAO subspecialties are Doctorate of Medicine (DM) courses that can be pursued after PG in Internal Medicine and include: Cardiology, Neurology, Nephrology, Gastroenterology, Endocrinology, Rheumatology, Pulmonary Medicine, Oncology, Hematology (can be pursued after Pathology or Internal Medicine), Neonatology (can be pursued after Pediatrics), Clinical Pharmacology (can be pursued after Pharmacology or Internal Medicine).

### Outcomes

The counts of the above-mentioned PG and SS SAO training spots were calculated. The SAO spot density was defined as the number of SAO training spots (PG and SS) per 10 million population. This was calculated to investigate population-based workforce production capacity. To investigate competitiveness in training, the proportion of PG SAO spots per 100 MBBS spots and the proportion of SS SAO spots per 100 PG SAO spots were assessed. For SS post per 100 PG spots under General Surgery and Otorhinolaryngology spots were considered as PG spots due to the reasons mentioned above. All outcomes (densities and proportions) were calculated for 36 states/union territories and the above-mentioned SAO specialties and subspecialties.

### Statistical Analysis

Wilcoxon’s signed rank test was used to test if SAO and non-SAO PG and SS spot densities differed significantly at a significance level set at alpha=5%. Data was wrangled using Google Sheets, analyzed in JASP v0.16.0.0, and visualized using Datawrapper.

## Results

### Postgraduate (PG) and Subspecialty (SS) SAO Spot Counts

In 2018, nationally, there were a total of 30087 postgraduate (PG) training spots in India. Of these, 13793 (45.84%) belonged to SAO specialties. Of the PG SAO spots, 2165 were diploma spots and 11628 were Masters in Surgery (MS) spots. Further distribution of PG SAO spots showed more General Surgery (3107), OBGYN (2842), and Anesthesia (3272) spots compared to Orthopedics (1930), Ophthalmology (1577), and Otorhinolaryngology (1065). Of the total 2494, 1305 (52.32%) sub-specialty (SS) training spots were SAO.

Genitourinary Surgery/Urology (250), Neurosurgery (245), Plastic Surgery (201), Cardiothoracic Surgery (192) and Pediatric Surgery (167) formed the major proportion of SS SAO spots whereas the rest of the SAO sub-specialty spots summed up to only 250 **(Figures 1A&B)**.

**Figure 1:**
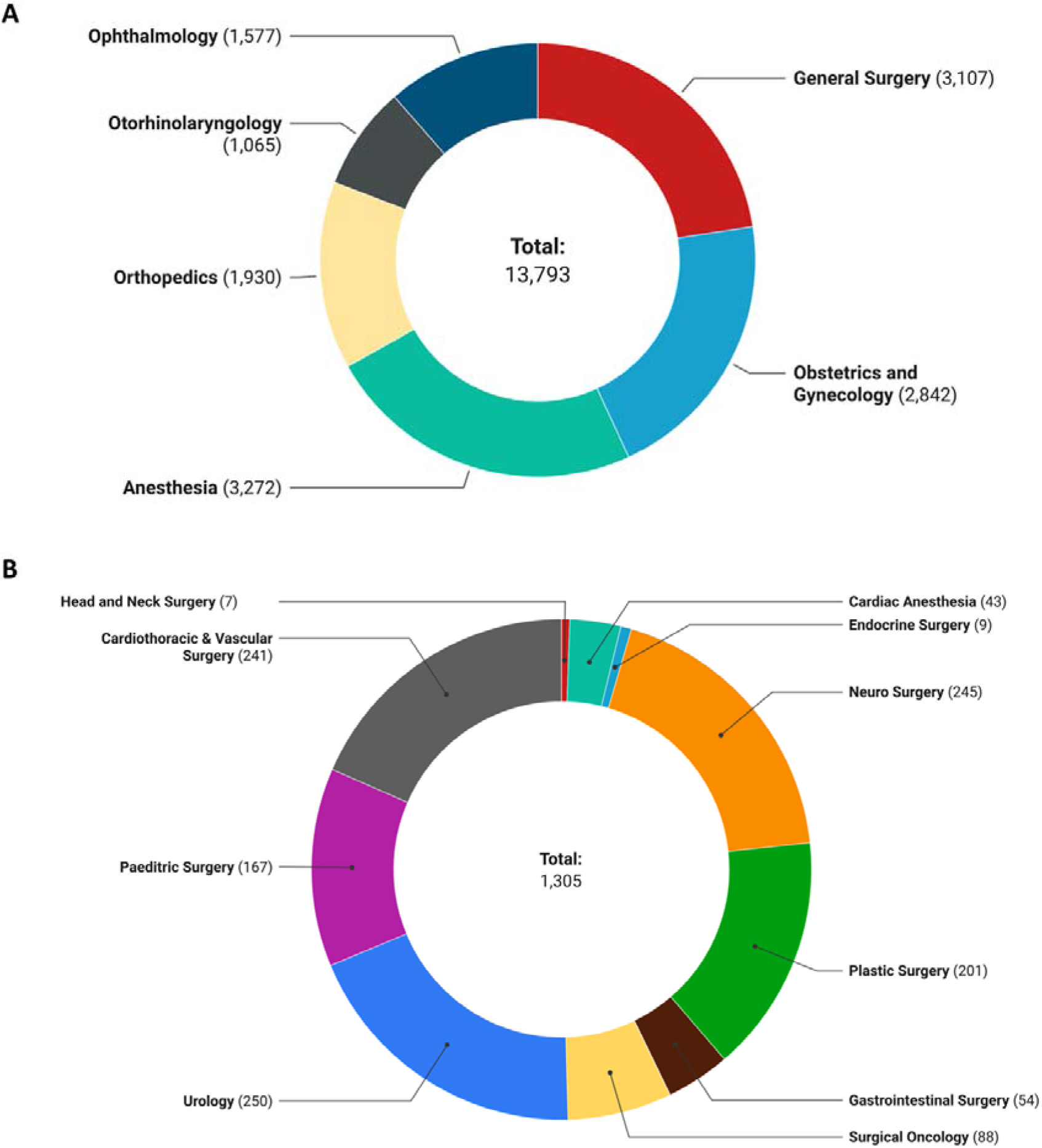
Distribution of surgery, anesthesia, and obstetrics related A) postgraduate and B) subspecialty training spots (counts).

### State-wise PG and SS SAO Spot Densities

The national PG & SS SAO spot densities were found to be 104.60 and 9.90 per 10 million population respectively. The distribution of PG & SS SAO spots densities was uneven with about two-thirds of spots concentrated in only 7 states and union territories including Chandigarh (PG SAO spot density: 1772.69, SS SAO spot density: 441.05), Puducherry (1619.047, 156.46), and Delhi (424.95, 96.40). Bihar (2.61, 0.08), Jharkhand (19.26, 0.27), Chhattisgarh (19.41, 0), Telangana (20.37, 1.95) and Uttar Pradesh (42.83, 3.20) had very low SAO spot densities. There were zero PG and SS SAO spots available in three northeastern states and four union territories **(Figures 2A&B)**. The PG SAO spot density was significantly lower compared to non-SAO spot density across states (n=36, effect size=−0.949, *p*<0.001) For state-level paired differences see **Figure 3A**. While the SS SAO spot density was higher than the non-SAO spot density across states, it was not statistically significant (n=36, effect size=0.287, *p*=0.197). For state-level paired differences see **Figure 3B**.

**Figure 2:**
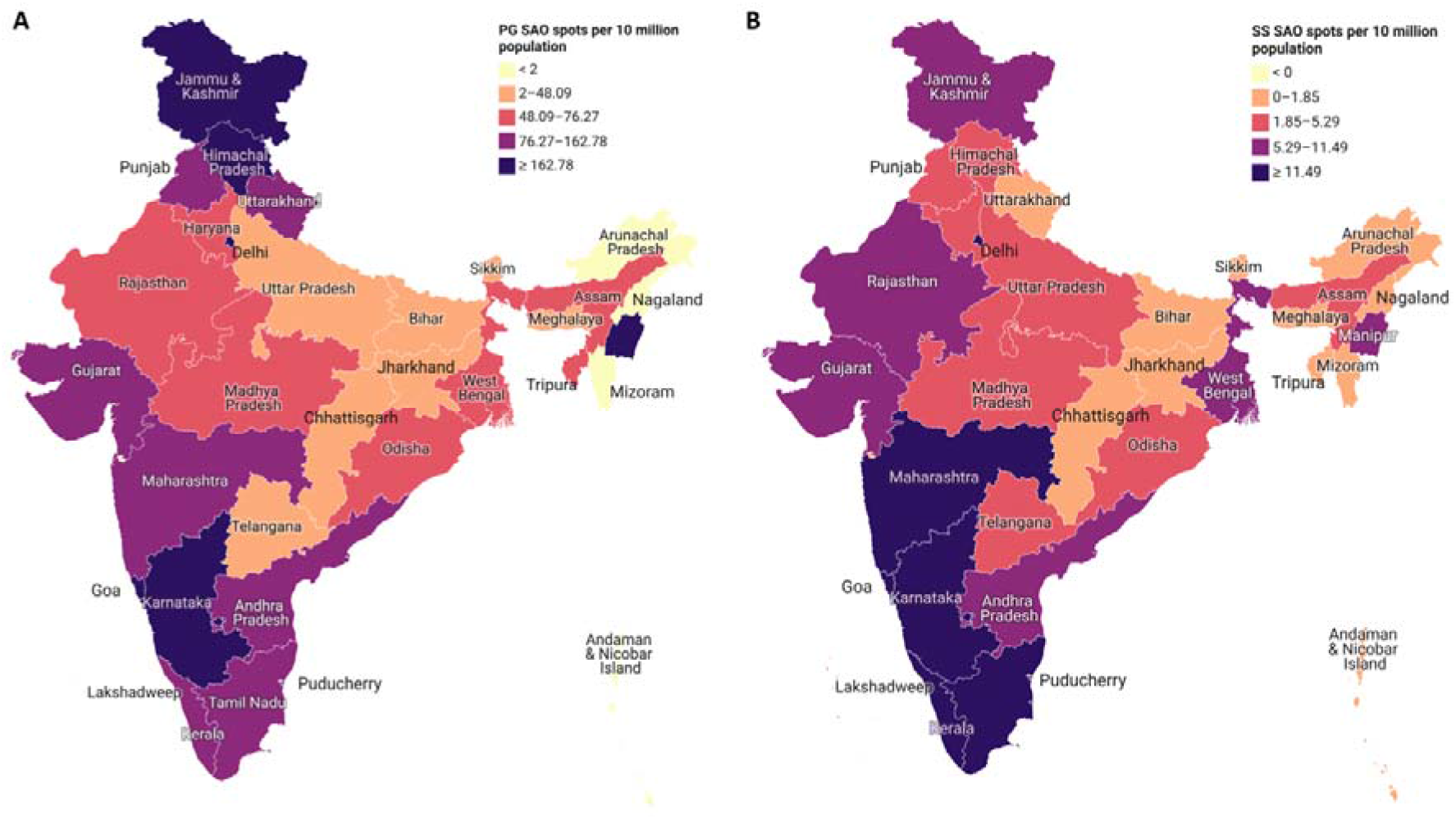
Densities per 10 million people across states and union territories of A) postgraduate and B) subspecialty surgery, anesthesia, and obstetrics training spots. All Diploma and MD/MS programs spots in General Surgery, Obstetrics & Gynecology, Anesthesia, Orthopedics, Ophthalmology and Otorhinolaryngology have been included in PG SAO spots. All MCh program spots in Urology (or Genitourinary Surgery), Neurosurgery, Plastic Surgery, Cardiothoracic and Vascular Surgery (including spots specific to Vascular Surgery and Thoracic Surgery), Pediatric Surgery, Onco-surgery, Gastrointestinal Surgery, Endocrine Surgery, Head-Neck Surgery and DM Cardiac Anesthesia have been included in SS SAO spots. Both maps use quantile breaks.

**Figure 3:**
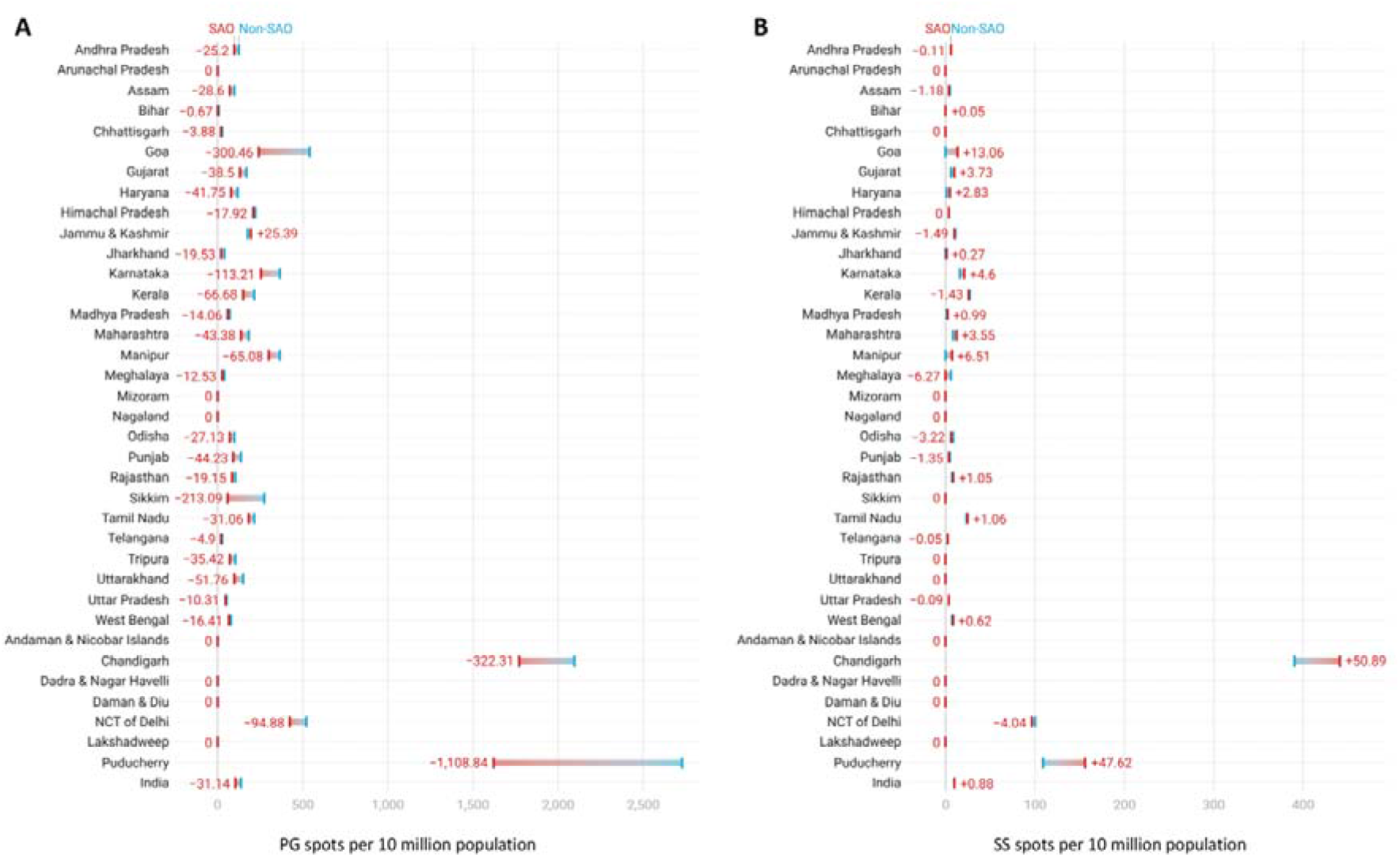
Comparison of surgery, anesthesia, and obstetric (SAO) and non-SAO spot densities for A) Postgraduate and B) Subspecialty training programs across states and union territories. SAO is depicted by red while non-SAO is depicted by blue. Difference is calculated as SAO spot density - non-SAO spot density. Negative differences depict lower SAO spot densities than non-SAO spot densities. States have been arranged in alphabetical order.

### Specialty-wise PG and SS SAO Spot Densities

Specialty-wise distribution of PG SAO spot densities (spots per 10 million population) for India and states is depicted in **Figure 4**. At the national level spot densities for General Surgery (23.56), Anesthesia (24.81), and OBGYN (21.55) were comparable and were much higher compared to Orthopedics (14.64), Ophthalmology (11.96), and Otorhinolaryngology (8.08). The lowest SAO spot density was observed in Bihar (Surgery=0.95, Anesthesia=0.35, OBGYN=0.48) while the highest was seen in Chandigarh (Surgery=780.32, Anesthesia=483.46, OBGYN=245.97). As depicted in **Figure 5**, national-level specialty-wise SS SAO spot densities were greater for Urology (1.90) and Neurosurgery (1.86) by Cardiothoracic and Vascular Surgery (1.83), followed by Plastic Surgery (1.52), and Pediatric Surgery (1.27). Spot densities of other super specialties fell below 1 per 10 million population.

**Figure 4:**
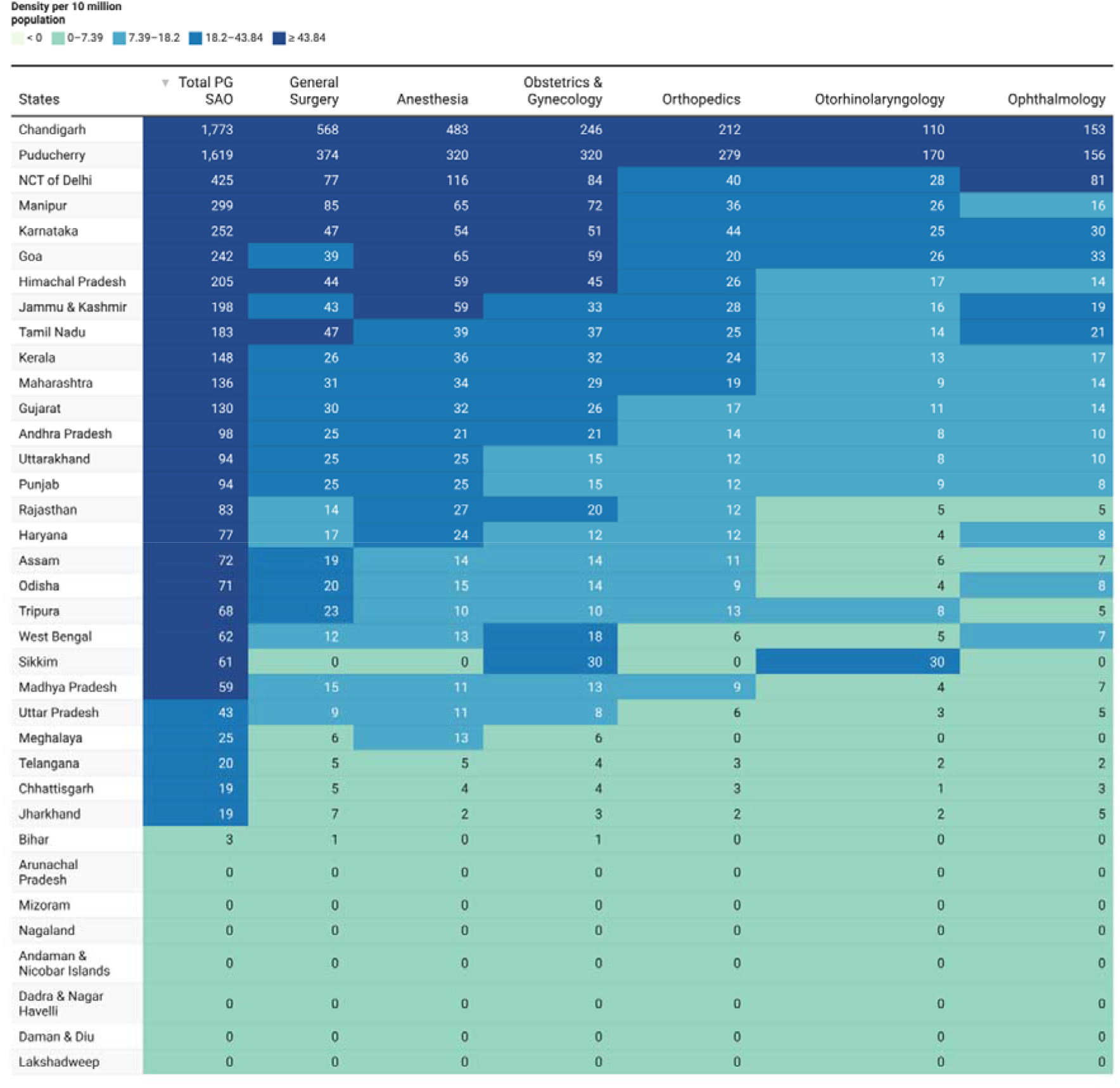
State-wise surgery, anesthesia, and obstetrics postgraduate spot density (per 10 million population) across specialties. Heatmap arranged in descending order of spot densities.

**Figure 5:**
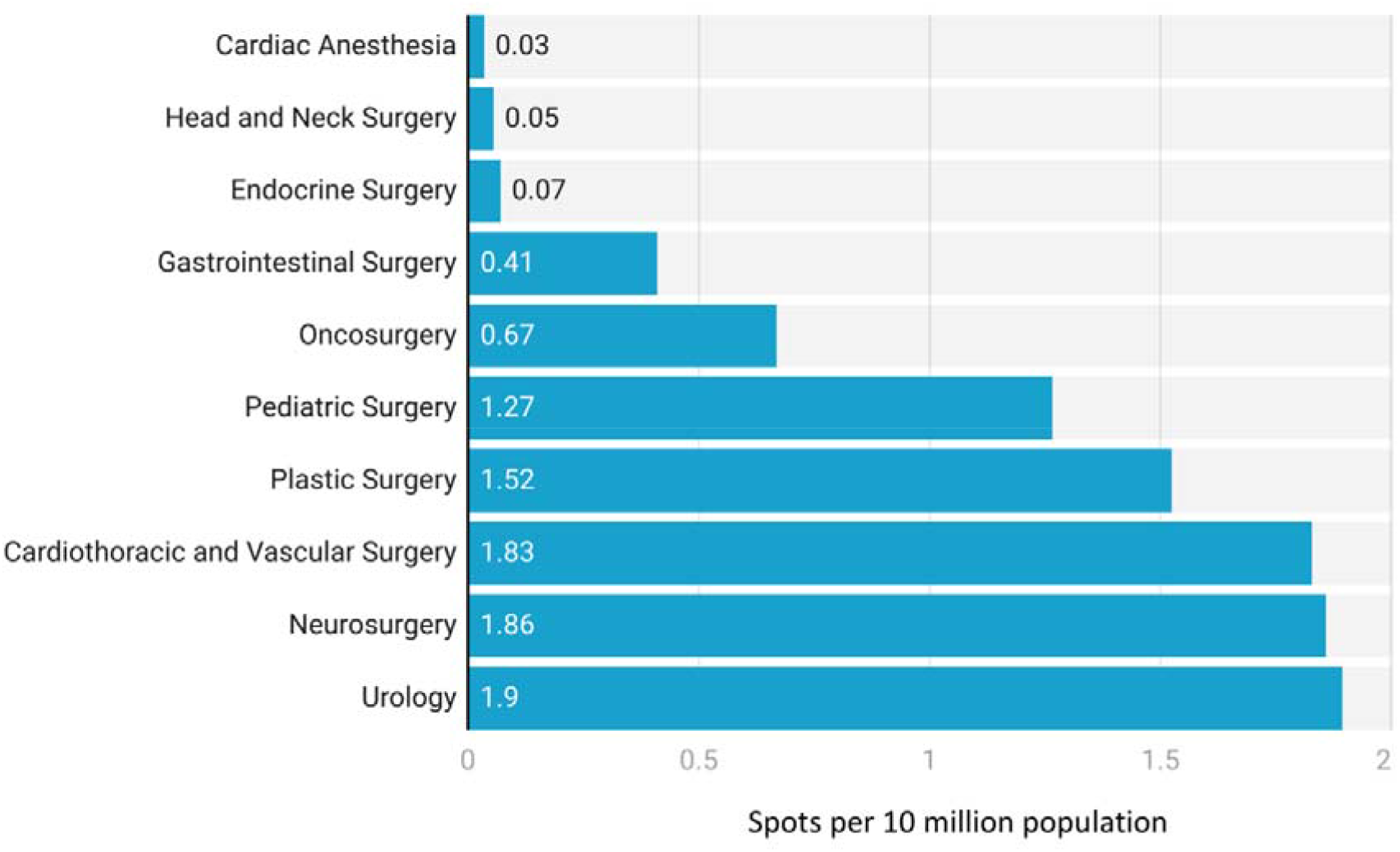
National surgery, anesthesia, and obstetrics subspecialty training spot density (per 10 million population) across specialties.

### Competitiveness of matching into SAO specialties and subspecialties

With 69405 undergraduate (MBBS) spots, there were 20 PG SAO spots per 100 MBBS spots. These proportions were 4:100 for General Surgery, OBGYN, and Anesthesia, 3:100 for Orthopedics, 2:100 for Ophthalmology, and 2:100 for Otorhinolaryngology. There were 40 SS SAO spots per 100 PG SAO spots. The proportions were 8:100 for Urology and Neurosurgery, 8:100 for Cardiothoracic and Vascular Surgery, 6:100 for Plastic Surgery, 5:100 for Pediatric Surgery, 3:100 for Onco-surgery, and 2:100 for Gastrointestinal Surgery. The state-wise PG to MBBS and SS to PG spot proportions are depicted in **Figure 6**. Chandigarh had the highest PG SAO spots per 100 MBBS seats and Delhi had the highest SS SAO spots per 100 PG SAO spots. Goa, Maharashtra, West Bengal, Tamil Nadu, Gujrat, Rajasthan, Odisha, Telangana, Karnataka, Andhra Pradesh, and union territories of Delhi and Puducherry had SS to PG proportions higher than PG to MBBS proportions.

**Figure 6:**
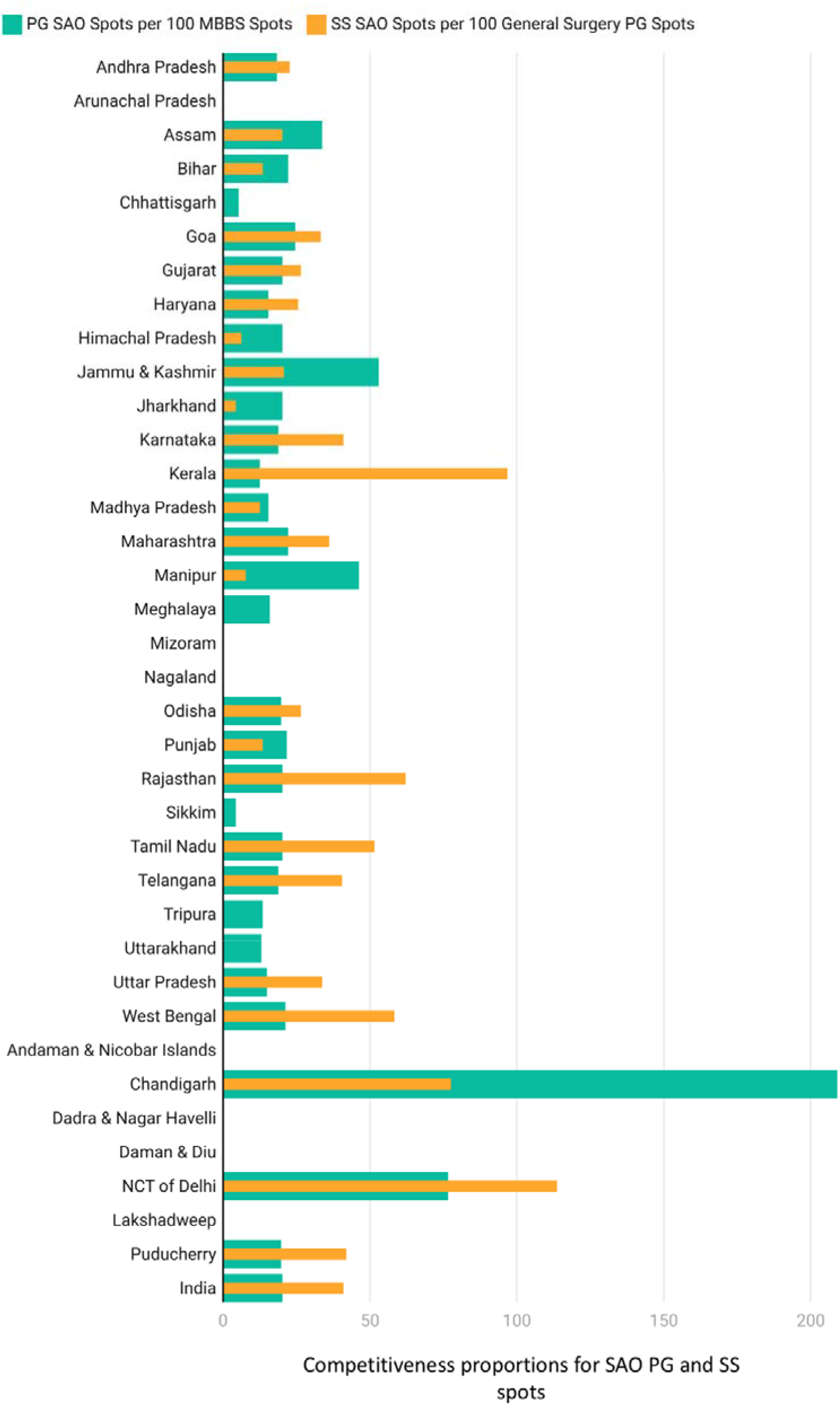
Competitiveness of matching into SAO PG (per 100 MBBS spots) and SS (per 100 SAO PG) spots across states and union territories. Footnotes: MBBS = Bachelor of Medicine and Bachelor of Surgery, PG = Postgraduate, SS = Subspecialty, SAO = Surgery, Anesthesia, and Obstetrics

## Discussion

### Summary of Findings

Nationally, SAO training spots form less than half of the total number of postgraduate (PG) spots. Moreover, General Surgery training spots formed only a tenth of the total PG spots. The spot counts, densities relative to populations, and proportions per 100 MBBS spots for General Surgery, Anesthesia, and OBGYN were similar to each other and almost twice that of Orthopedic Surgery, Ophthalmology, and Otorhinolaryngology. For subspecialty (SS) spots, more than half belonged to SAO subspecialties. The spot counts, densities, and proportions for Urology, Neurosurgery, and Cardiothoracic and Vascular Surgery were greater than those for the rest of the SAO subspecialties.

Annually, India can produce only about 24 General Surgeons, 15 Orthopedic Surgeons, 12 Ophthalmologists, 8 Otorhinolaryngologists, 25 Anesthetists, 22 OBGYNs, and 10 sub-specialist surgeons per 10 million population. There is disparity across states for spot densities with Bihar, Jharkhand, Chhattisgarh, Telangana, and Uttar Pradesh having severely limited SAO workforce production capacity. Spot density is a normalized parameter to look at equitable distribution of spots and hence is preferred over absolute spot counts for comparison across states. In terms of competitiveness to get the SAO specialties, for every 100 MBBS graduates, only four can get into either General Surgery, Anesthesia, or OBGYN, three can get into Orthopedic Surgery, and only two can get into Ophthalmology or Otorhinolaryngology. However, compared to PG, competitiveness is lower to get into SS subspecialties such as Urology, Neurosurgery, and Cardiothoracic and Vascular Surgery.

### Interpretation and Caveats

The above-mentioned numbers about the SAO training spots are the maximum (or cap) number of available spots. Hence, the current analysis is about production capacity and not the actual SAO trainee numbers or sociodemographic characteristics (gender, age, etc.) of the trainees. The current analysis also does not attempt to investigate the adequacy and quality of training that are certainly critical for ensuring competent SAO workforce. It is important to note that in some instances, available spots may not correspond to the actual number of trainees who get admitted into particular specialty programs every year. In recent times in India, there has been a trend towards MBBS graduates preferring non-SAO specialties like Internal Medicine, Dermatology, and Radiology over SAO branches like General Surgery, OBGYN, etc.^13^ Hence, SAO spots in some states go vacant every year, especially for surgical subspecialties including Cardiothoracic and Vascular Surgery, Plastic Surgery, Pediatric Surgery, and Neurosurgery.^14,15^ There are multiple reasons underlying graduates’ preferences and thereby vacancies. The specialty-specific issues include availability of resources in hospitals and supportive surgical teams, work-life balance and need to attend fewer emergencies, among others.^14,16,17^ Another reason is the limited practical exposure to SAO specialties in terms of attending or performing procedures during the MBBS curriculum and internships.^18^ Vacancies are also determined by state policies. For instance, the policy of compulsory government service (bond) for a fixed period of years (1-5 years) after post-training influences the graduates’ choice to get admitted to PG or SS programs in a given state.^19^ States with stringent and longer bonds are less preferred by graduates, leading to counterproductive migration of the SAO personnel. This worsens the existing disparities in the workforce right at the training level. Major barriers preventing young trainees from opting surgical subspecialties include poor patient prognosis, few number of cases and limited scope of practice.^15^

### Contextualizing Findings

Previously, there has been only one other study looking at SAO workforce production capacity in India.^10^ Compared to the WHO sponsored study to estimate the workforce production, there has been an increase of PG SAO spots by about 52%, from 9048 in 2015 to 13793 in 2018. The PG SAO spot density has increased by about 40% from 75 to 105 per 10 million population. Among states, the previous study only investigated Kerala whereas the current study included the workforce capacity across 36 states and union territories. The SAO spots in Kerala have increased from 412 to 518 (25.7%) from 2015 to 2018 while the spot density has increased from 123 to 148 (20.3%). Comparable data from other low- and middle-income countries is scant. However, a retrospective longitudinal (2014-18) review from the United States found that in 2018, there were 41.8 ± 5.2 General Surgery residency spots per 10 million population.^20^ This is almost twice as compared to India’s General surgery spot density of 23.56.

The SAO workforce projection study conducted after the LCoGS noted that India needs to train 291,824 SAO personnel overall to reach the LCoGS target of 20 SAO personnel per 100,000 people by 2030.^4^ Briefly, assuming a linear growth starting from 2021 (i.e., year of graduation for those entering in 2018), at the current production capacity of 13793 SAO PG graduates per year, India can produce 124,137 new PG SAO personnel by 2030. Not accounting for attrition or assuming any differences in growth rates, this still depicts that production capacity needs massive enhancement. However, more nuanced modeling studies that involve state-level differences, vacancy rates, attrition in workforce due to death and retirement, attrition in training, etc. are needed to create accurate estimates on the extent to which training spots should be increased.

### Policy Implications

SAO care needs to be an integral part of the public health plans and policies in India.^21^ Such planning would require investigating the SAO workforce situation in India.^22,23^ To meet the LCoGS target, there is a need to scale up SAO workforce, which in turn depends on increase in the number of SAO training spots. Medical education policies need to give particular attention to Central, Northern, and Northeastern states with poor spot densities to achieve equitable distribution. Spots of PG SAO specialties and subspecialties need to be further assessed as per the state-level disease burdens relevant to them. Currently, there is no indication that policies account for population needs including disease burdens. For instance, the density of Ophthalmology training spots should be based on local needs including state-level disease burden and proportion of the aging population that are at risk for cataracts and glaucoma.^24,25^ Similarly, there needs to be a greater focus on scaling up Pediatric Surgery SS training spots given the large pediatric population along with high neonatal and infant mortality rate in India.^26,27^

### Strengths and Limitations

To our knowledge, this is the first pan-India analysis of postgraduate and subspecialty spot distribution with special emphasis on SAO specialties across 36 states and union territories. Our outcome variables, spot density relative to population and spot proportions are easily calculable normative variables to assess the workforce production capacity and competitiveness, respectively. These can be used across regions and times periods for consistent comparisons.

However, this study has several limitations. First, the study used 2018 data. Most likely, there have been improvements in the training spot counts in the last four years. However, we had to rely on 2018 data as it is the most recent systematic state-level data available. Though a notification about increase in training spots has been issued, the updated data is not currently available.^28^ Second, private hospitals that are not affiliated to a medical college provide a Diplomate of National Board (DNB) degree which is equivalent to an MS degree. Data on DNB spots was unavailable and has not been included in this study. However, generally DNB spots (SAO and non-SAO) form less than 9% of total PG spots.^29,30^ Third, we could only provide state level data and do not account for differences within states at district levels or those between urban and rural regions. Since most of the medical colleges are concentrated in a few cities, district level data would provide a better understating and intra-state disparities in SAO workforce production. Fourth, the current analysis does not account for paramedical staff valuable to SAO care including operation theater nurses or trained midwives who are also a part of the surgical workforce. Finally, due to lack of data, we do not investigate the distribution of training spots as per affirmative action or reservation policies based on caste groups and socioeconomic classes.

## Conclusion

India’s SAO workforce production capacity needs to be scaled up for the SAO workforce to meet the LCoGS target by 2030. The distribution of PG and SS SAO training spots is inequitable across the states as identified by the differences in the spot densities. Policies need to focus on specific states and SAO specialties that urgently require scale up to achieve equitable distribution and meet population needs. Future studies can build on the current findings to estimate the required rate of scale up of production capacity.

## Data Availability

All data produced in the present study are available upon reasonable request to the authors

## Abbreviations

SAO: Surgical, Anesthesia and Obstetrics
PG: Postgraduate
SS: Subspecialty
MBBS: Bachelor of Medicine and Bachelor of Surgery
MS: Master of Surgery
MD: Doctor of Medicine
MCh: Master of Chirurgiae
DM: Doctorate of Medicine
NEET: National Eligibility cum Entrance Test
OBGYN: Obstetrics & Gynecology
LCoGS: Lancet Commission on Global Surgery

## Author Contributions

### Shirish Rao

Conceptualization, Methodology, Formal Analysis, Data Curation, Writing - Original Draft, Review & Editing.

### Siddhesh Zadey

Conceptualization, Methodology, Writing - Review & Editing, Supervision, Project Administration.

## Acknowledgements

We thank Tejali Gangane and Ram Pachpor for helping with data extraction and Dr. Anveshi Nayan for feedback on the manuscript.

## Ethics approval and consent to participate

Not applicable

## Consent for publication

Not applicable

## Availability of data and materials

All data used in the manuscript can be requested from the authors.

## References

1. Karan A, Negandhi H, Nair R, Sharma A, Tiwari R, Zodpey S. Size, composition and distribution of human resource for health in India: new estimates using National Sample Survey and Registry data. BMJ Open. 2019;9(4):e025979. doi:10.1136/bmjopen-2018-025979

2. GBD 2019 Human Resources for Health Collaborators. Measuring the availability of human resources for health and its relationship to universal health coverage for 204 countries and territories from 1990 to 2019: a systematic analysis for the Global Burden of Disease Study 2019. Lancet. 2022;399(10341):2129–2154. doi:10.1016/S0140-6736(22)00532-3

3. Holmer H, Lantz A, Kunjumen T, et al. Global distribution of surgeons, anaesthesiologists, and obstetricians. Lancet Glob Health. 2015;3 Suppl 2:Supplementary Appendix Table S4. doi:10.1016/S2214-109X(14)70349-3

4. Daniels KM, Riesel JN, Verguet S, Meara JG, Shrime MG. The Scale-Up of the Global Surgical Workforce: Can Estimates be Achieved by 2030? World J Surg. 2020;44(4):1053–1061. doi:10.1007/s00268-019-05329-9

5. National Medical Commission. UG Curriculum. 2019. Accessed January 10, 2023. https://www.nmc.org.in/information-desk/for-colleges/ug-curriculum/

6. National Medical Commission. PG Curricula. 2019. Accessed January 10, 2023. https://www.nmc.org.in/information-desk/for-colleges/pg-curricula-2/

7. NBEMS. NEET-PG. 2019. Accessed January 10, 2023. https://natboard.edu.in/viewnbeexam?exam=neetpg

8. NBEMS. NEET-SS. 2019. Accessed January 10, 2023. https://natboard.edu.in/viewnbeexam?exam=neetss

9. Jain G, Are C, Agrawal V, Agarwal P. General surgery training in the USA, UK, and india: a scrutiny of strength and challenges. Indian J Surg. 2022;84(S1):318–325. doi:10.1007/s12262-021-03226-x

10. Country Office for India, World Health Organization. Surgical Workforce in India: What the State of Kerala Tells Us about the Production, Stock and Migration of the Health Workforce. (Rao K, Arora RA, Bhatnagar A, et al., eds.). WHO; 2015. Accessed July 29, 2022. https://apps.who.int/iris/handle/10665/246163

11. Central Bureau of Health Intelligence (CBHI). National Health Profile (NHP) of India - 2019. Ministry of Health and Family Welfare; 2019.

12. National Commission on Population. Population Projections for India and States 2011-2036. National Commission on Population, Ministry of Health and Family Welfare; 2019.

13. Chawla J, Arora M, Datta K, Singh SP, Arora A. Factors affecting the choice of postgraduate specialty among undergraduate medical students: a prospective observational study from India. SE Asian Jnl Med Educ. 2018;12(2):35. doi:10.4038/seajme.v12i2.50

14. Chhapia H. Vacant Seats In Courses For Non-surgical Branches Too. Times of India. July 31, 2022. Accessed January 10, 2023. https://timesofindia.indiatimes.com/city/mumbai/vacant-seats-in-courses-for-non-surgical-branches-too/articleshow/93244787.cms

15. Vatyam N. Telangana: No takers for 40 super specialty seats despite zero percentile rider. Times of India. https://timesofindia.indiatimes.com/city/hyderabad/no-takers-for-40-super-speciality-seats-despite-zero-percentile-rider/articleshow/93260479.cms. Published August 1, 2022. Accessed December 28, 2022.

16. Sharma JP. Medical colleges: 352 vacant super speciality seats cost exchequer Rs 200 crore. Hindustan Times. November 20, 2017. Accessed January 10, 2023. https://www.hindustantimes.com/india-news/352-vacant-super-speciality-medical-seats-cost-exchequer-rs-200-crore/story-rP0OZKBDMonLyn8Ba2oMhK.html

17. Mishra B. More than 300 Clinical Seats Vacant: Doctors demand special round of NEET PG Counselling. Medical Dialogues. December 21, 2022. Accessed January 10, 2023. https://medicaldialogues.in/amp/news/education/more-than-300-clinical-seats-vacant-doctors-demand-special-round-of-neet-pg-counselling-104380

18. Are C. Surgical training in india versus abroad: what more needs to be done? Indian J Surg Oncol. 2013;4(4):382. doi:10.1007/s13193-013-0266-3

19. PTI. Health Ministry working on guidelines to scrap bond policy for doctors. The Hindu. https://www.thehindu.com/news/national/health-ministry-working-on-guidelines-to-scrap-bond-policy-for-doctors/article66104090.ece. Published 2022. Accessed January 11, 2023.

20. Elkbuli A, Narvel RI, Dowd B, McKenney M, Boneva D. Distribution of general surgery residencies in the united states and gender inequality: are we there yet? J Surg Educ. 2019;76(6):1460–1468. doi:10.1016/j.jsurg.2019.05.008

21. Zadey S, Sonal S, Iyer H, et al. Roadblocks and solutions to planning surgical care for a billion Indians. BMJ Glob Health. 2022;7(11). doi:10.1136/bmjgh-2022-010292

22. Zadey S, Iyer H, Nayan A, et al. Evaluating the Status of the Lancet Commission on Global Surgery Indicators for India. Published online 2022. Accessed October 25, 2022. https://papers.ssrn.com/sol3/papers.cfm?abstract_id=4242033

23. Zadey S. Why India’s Surgical Care Crisis Is Less Jigsaw, More Tetris – in Three Charts. The Wire. https://thewire.in/health/why-indias-surgical-care-crisis-is-less-jigsaw-more-tetris-in-three-charts. Published October 12, 2019. Accessed August 28, 2021.

24. Pandey S, Sharma V. Ophthalmology training and teaching in India: How these young ophthalmologists can become leaders of tomorrow? Indian J Ophthalmol. 2018;66(10):1517. doi:10.4103/ijo.IJO_898_18

25. Das T, Panda L. Imagining eye care in India (2018 Lalit Prakash Agarwal lecture). Indian J Ophthalmol. 2018;66(11):1532. doi:10.4103/ijo.IJO_872_18

26. Shah R. The past, the present, and the future of pediatric surgery in India. J Indian Assoc Pediatr Surg. 2015;20(1):2–7. doi:10.4103/0971-9261.145436

27. Gupta DK. Pediatric surgery in India: Time now for review. J Indian Assoc Pediatr Surg. 2015;20(2):57–59. doi:10.4103/0971-9261.151543

28. Ministry of Health and Family Welfare. Governance Reforms in Medical Education (2014-2022). 2022. Accessed January 10, 2023. https://medicaldialogues.in/pdf_upload/education-booklet-191670.pdf

29. National Board of Examination. NBE Accredited Seats. 2022. Accessed January 9, 2023. https://accr.natboard.edu.in/online_user/frontpage.php?v=4

30. National Medical Commission. Dashboard of Medical Faculty & PG Students in India. 2020. Accessed January 9, 2023. https://www.nmc.org.in/information-desk/faculty-medical-students-information/

